# D-Cycloserine for Treatment of Chronic Low Back Pain: Results from a Randomized, Double-Blind, Placebo-Controlled Clinical Trial

**DOI:** 10.1101/2025.09.14.25335705

**Authors:** Joana Barroso, Andrew D. Vigotsky, Santiago Espinosa Salas, Olivia Cong, Narina Simonan, A. Vania Apkarian, Thomas Schnitzer

## Abstract

**Objective:** Chronic low back pain (cLBP) is a leading cause of global disability, with current treatments offering limited and inconsistent relief. D-cycloserine (DCS), a partial NMDA receptor agonist and FDA-approved antimicrobial, has shown promise in preclinical models for reducing neuropathic pain and its negative emotional impact. Building on these findings and a positive pilot study, we conducted a randomized, double-blind, placebo-controlled phase 2 trial to evaluate the efficacy and safety of DCS in adults with cLBP.

**Methods:** Participants with cLBP (n=203) were randomized to receive 200 mg DCS or placebo twice daily for 12 weeks, followed by a 12-week placebo phase. The primary outcome was pain intensity (numeric rating scale, NRS) at 12 weeks; secondary outcomes included pain intensity at 24 weeks, safety, and patient-reported psychological measures.

**Results:** DCS had negligible effects on pain compared to placebo at both 12 weeks (adjusted effect: −0.07/10 NRS units, 95% CI: (−0.67, 0.53), *p* = 0.78) and at 24 weeks (adjusted effect: −0.22/10 NRS units, 95% CI: (−0.84, 0.40), *p* = 0.39). Adverse events were similar between groups. Exploratory outcomes also showed no significant effects.

**Conclusion:** Our results indicate that 12 weeks of DCS 400 mg/day does not provide clinically meaningful analgesia in cLBP, though it is well tolerated. These findings highlight the challenges of translating preclinical neuropathic pain results to heterogeneous clinical pain populations.

## 1. Introduction

Low back pain (LBP) represents a significant global health challenge, affecting over 619 million individuals worldwide in 2020, with projections indicating this number will surge to 843 million by 2050^1^. As a leading cause of global disability, LBP profoundly impacts individual well-being, physical function, and quality of life^2-4^. It also has a substantial impact on healthcare expenditure, and in the US alone, reached approximately $134.5 billion in 2016 (for low back and neck pain), surpassing spending on other major conditions^5^. Current treatments for chronic low back pain (cLBP) generally provide only small to moderate benefits^6^. Many cLBP patients receive multiple treatments with limited success, and many continue to experience pain or disability after completing or terminating medical treatments^7^. Moreover, although opioid analgesics are no more effective than non-opioid treatments for cLBP^8, 9^, opioid prescriptions remain common^10^, fueling the ongoing opioid crisis and emphasizing the urgent need for more effective, non-addictive therapies.

D-cycloserine (DCS), an FDA-approved antimicrobial agent used as a second-line treatment for tuberculosis, has shown potential in treating chronic pain^11, 12^. DCS is a partial agonist at the glycine site of the NMDA (N-methyl-D-aspartate) receptor. Consequently, at lower doses, it enhances NMDA receptor function, whereas at higher doses, it may act as a functional NMDA receptor antagonist^13^. Preclinical and clinical studies have demonstrated that DCS facilitates fear extinction and cognitive flexibility by potentiating NMDA receptor signaling during extinction training, thereby promoting adaptive synaptic plasticity^14, 15^. These effects appear to be mediated via corticolimbic circuits, including the medial prefrontal cortex (mPFC) and amygdala, which are critical for emotional regulation and aversive learning^16, 17^. Given that chronic pain is posited to be a disorder of maladaptive learning and affective dysregulation, targeting these circuits with DCS offers a novel therapeutic strategy^12, 18^.

Consistent with this rationale, preclinical studies from our group have shown that DCS reduces pain-like behaviors in rodent models of chronic pain, including spared nerve injury and chemotherapy-induced neuropathy^12^. The effects were dose-dependent and increased with prolonged use and associated with persistent analgesia for over a month after discontinuation^12^. The analgesic effects of DCS appeared to be specific to the injured areas, without affecting normal pain sensitivity elsewhere. Importantly, these analgesic effects required chronic dosing, suggesting the effect of DCS relies on long-term adaptation rather than immediate analgesia. Following these experimental results, our group conducted a small proof-of-concept pilot study of DCS in individuals with cLBP. In this randomized, parallel-group design with a placebo control, the DCS group showed a greater reduction in pain than the placebo group after 6 weeks of treatment^19^. Secondary analyses exploring psychological dimensions of pain revealed that DCS may positively influence emotional well-being^19^.

Building on compelling preclinical evidence and encouraging results from our initial human pilot study, we conducted a larger phase 2 study of DCS in patients with cLBP to investigate the efficacy and safety of DCS. The primary outcome measure of this study was the efficacy of pain reduction relative to placebo, assessed using the Numeric Rating Scale (NRS) pain ratings at the 12-week endpoint. Secondary outcomes included safety evaluation, longer-term pain relief (24 weeks), and a comprehensive set of patient-reported outcomes and psychological assessments, providing an evaluation of DCS’s impact on both pain intensity and associated psychological factors in cLBP management.

## 2. Methods

### 2.1 Study Design and Oversight

This parallel group, placebo-controlled, randomized controlled trial (RCT) was conducted at Northwestern University in Chicago, IL. The study protocol was approved by the Northwestern Institutional Review Board (STU00205398) and the study was registered on ClinicalTrials.gov (identifier: NCT03535688). The full trial protocol and statistical analysis plan are publicly accessible on the registry website. Deidentified individual participant data and supporting documents will be made available upon request to the corresponding author, contingent on a data use agreement.

All participants provided written informed consent before an initial screening to determine eligibility and interest. Participants recorded their pain levels twice daily using an electronic diary (eDiary) accessible via smartphone or computer. During the screening period, they discontinued medications used for cLBP and were provided acetaminophen (maximum dose 3000 mg/day) for pain relief. A baseline visit was scheduled after 10–14 days.

To qualify for randomization, participants needed a mean pain level of ≥4 and ≤9 over 5– 7 days prior to the baseline visit (with at least five eDiary entries during this period) and a pain level of ≥4 at baseline. At the baseline visit, participants completed questionnaires assessing pain, function, psychological factors, and quality of life, followed by a brain MRI (not reported in this paper). Eligible participants were then randomized to receive either 200 mg of DCS or a placebo, taken twice daily in identical capsules to ensure blinding.

Follow-up visits were conducted at 2, 6, 12, and 24 weeks after randomization. At each visit, participants completed questionnaires evaluating pain intensity and characteristics, and safety data were collected. Study medication was dispensed at the 2-week and 6-week visits. At the 12-week visit, participants underwent a second brain MRI along with regular assessments. Subsequently, all participants received placebo to evaluate the persistence of treatment effects over the next 12 weeks. Throughout the study, participants continued to record pain levels via the eDiary and were provided with rescue acetaminophen to take as needed.

### 2.2 Participants

Participants were recruited from two primary sources: Northwestern Medicine (NM) and the Shirley Ryan AbilityLab (SRALab) healthcare system, as well as the broader Chicago community. Recruitment from the Chicago community was supported through social media platforms like Facebook and third-party vendors such as *ClinicalConnection*. These combined approaches successfully facilitated participant enrollment for the trial.

Eligible participants for this study were adults aged 18 years or older with a history of low back pain lasting at least six months. Participants were required to be in generally stable health, willing to discontinue all other pain medications for chronic back pain, and abstain from alcohol during the study. Women of childbearing potential had to use effective contraception or be postmenopausal for at least one year. All participants were required to provide informed consent, agree to record their pain levels and treatment adherence via an eDiary, and comply with the study protocols.

Exclusion criteria included systemic symptoms such as fever, evidence of rheumatoid arthritis or other specific spinal conditions, recent back surgery, epidural steroid injections within the last three months, or litigation or disability claims related to back pain. Individuals with a history of seizures, significant psychiatric disorders, substance abuse, abnormal lab results, or medical conditions such as severe renal or cardiac disease were excluded. Participants using the following medications: ethionamide, dilantin, isoniazid medications, recreational drugs, or medical marijuana, or those with intra-axial implants, were not eligible. Pregnancy, breastfeeding, or inability to use effective contraception also disqualified participants. Additionally, any recent changes in medication or physical therapy regimens for back pain, as well as participation in other clinical trials within the last 90 days, were exclusionary.

### 2.3 Randomization

Participants were randomized in a 1:1 ratio to receive either DCS or placebo using a computer-generated permuted block randomization scheme with varying block sizes (2 and 4); randomization was performed on April 4, 2018, by an independent investigator not involved in other aspects of the study. Treatment allocation was concealed using sequentially numbered containers. Study medications were provided in identical opaque capsules to maintain double blinding. A designated unblinded staff member, who had no involvement in clinical assessments, was responsible for medication assignment and maintained the randomization code. This code was only accessible for documented emergencies requiring treatment unblinding after consultation with the site and principal investigator. The randomization codes were made available to study statisticians after the database lock at study conclusion.

### 2.4 Outcomes and Assessments

The primary outcome measure of this study was the efficacy of pain reduction, assessed using the numeric rating scale (NRS). Specifically, the mean NRS scores for the 7 days preceding the 12-week endpoint in the DCS group were compared to those in the placebo group, adjusted for baseline pain (mean NRS score from the 7 days preceding randomization). Pain was measured using an 11-point NRS scale.

Secondary outcome measures included pain at 24 week, evaluated between weeks 12 and 24 after the discontinuation of DCS therapy. Exploratory outcomes including a variety of patient-reported outcomes and psychological assessments were collected at various time points to provide a comprehensive understanding of the treatment’s impact. These assessments included the Patient Global Assessment (PGA), the Patient Global Impression of Change (PGIC), and the McGill Pain Questionnaire (MPQ), which were administered at screening, Week 12, and Week 24. Similarly, the painDETECT instrument, Beck Depression Inventory (BDI) – Second Version, Positive and Negative Affect Schedule (PANAS), Pain Catastrophizing Scale (PCS), and other functional and psychological measures, including the Oswestry Disability Index (ODI) and SF-12 Health Survey, were collected at screening, Week 12, and Week 24.

(add refs)

### 2.5 Protocol Deviation in Treatment Assignment

Study treatment allocation was in a 1:1 ratio of DCS to placebo in permuted blocks of 2 and 4. After the first 170 participants were entered into the study, errors in study drug allocation occurred such that many participants did not receive their appropriately allocated treatment. Therefore, all efficacy analyses were conducted using only the first 170 participants who received drug based on the correct allocation assignment. The safety analyses included all participants, with participants being assigned to the treatment that they actually received.

### 2.6 Statistical Analysis

The sample size for this study was determined using data from a prior pilot trial^19^; based on this data, our expected effect size was estimated at Cohen’s d = 0.4 between the treatment and placebo groups. To achieve 80% statistical power at a significance level of α = 0.05, it was calculated that 98 participants per group would be required. To account for an anticipated dropout rate of 20% at the primary endpoint of Week 12, the adjusted sample size increased to 122 participants per group, resulting in a total of 244 participants for the study. No interim analyses were planned or performed.

Baseline pain was calculated as the mean of all NRS ratings during the week preceding randomization, with no missing data due to mandatory compliance (≥ 5 ratings). Week 12 pain was defined as the mean of all ratings during that week.

Missing data were categorized into six mechanisms based on participants’ self-report: withdrawal of consent, lack of efficacy, loss to follow-up, adverse events, noncompliance, or early study termination. Multivariate imputation by chained equations (MICE) was employed, incorporating baseline pain, age, sex, and prior pain ratings. Participants in withdrawal, loss to follow-up, noncompliance, or early termination groups were imputed under missing-at-random assumptions using randomized group models. Those discontinuing due to lack of efficacy or adverse events were imputed under a “jump-to-reference” assumption using placebo group models. Imputation was performed separately for each treatment arm to account for potential interaction effects.

The primary analysis assessed the effect of DCS on Week 12 pain relative to placebo, adjusted for baseline pain, using an analysis of covariance (ANCOVA) estimated with ordinary least squares:

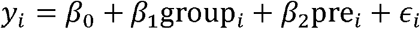

where *y*_*i*_ is Week 12 pain, group_*i*_ indicates treatment assignment (DCS=1, placebo=0), and pre_*i*_ is the baseline pain for subject *i*. Model fits were assessed by examining residual behavior. Effect estimates were reported with 95% confidence intervals (CI), and *p*-values for the primary outcomes were derived from a randomization test, in which randomizations were performed within each randomization block. As a secondary outcome, we examined the effect of DCS on pain ratings at Week 24 using the same model, 12 weeks after DCS participants transitioned to placebo. Finally, we estimated and contrasted the above treatment effects between males and females by including sex and group-by-sex terms in the model.

As a sensitivity analysis, we performed an as-treated analysis, in which all participants— including those who were misrandomized—were analyzed based on the treatment they received. Notably, this analysis is inconsistent with the randomization structure and ITT principles. Secondary analyses were conducted for the remaining outcomes, which were modeled using ANCOVAs (with baseline as a covariate) or ordinal regression. When model assumptions were clearly violated, heteroskedasticity was modeled using the HC3 sandwich estimator and departures from normality were addressed using permutations and the bias-corrected and accelerated bootstrapped CIs.

Analyses were conducted using R (version 4.0.0), with MICE (version 3.16.0) for imputation and concurve for consonance functions. All modifications and assumptions were transparently reported.

## 3. Results

### 3.1 Trial population and baseline characteristics

A total of 436 participants were screened, with 233 excluded, primarily for not meeting inclusion criteria (n=136). Of the 203 randomized participants, 102 were allocated to the DCS group and 101 to placebo. The efficacy population included 86 DCS and 84 placebo participants who received at least one dose of the intervention, while all 203 randomized participants were included in the safety analysis. By Week 12, 73 DCS and 76 placebo participants completed the primary endpoint assessment, decreasing to 57 and 62, respectively, by Week 24. Discontinuations were due to loss to follow-up, noncompliance, adverse events, or lack of efficacy. Notably, protocol deviations led to the exclusion of 27 DCS and 6 placebo participants from efficacy analyses but not safety analyses. **Figure 1** shows participant flow details.

**Figure 1.**
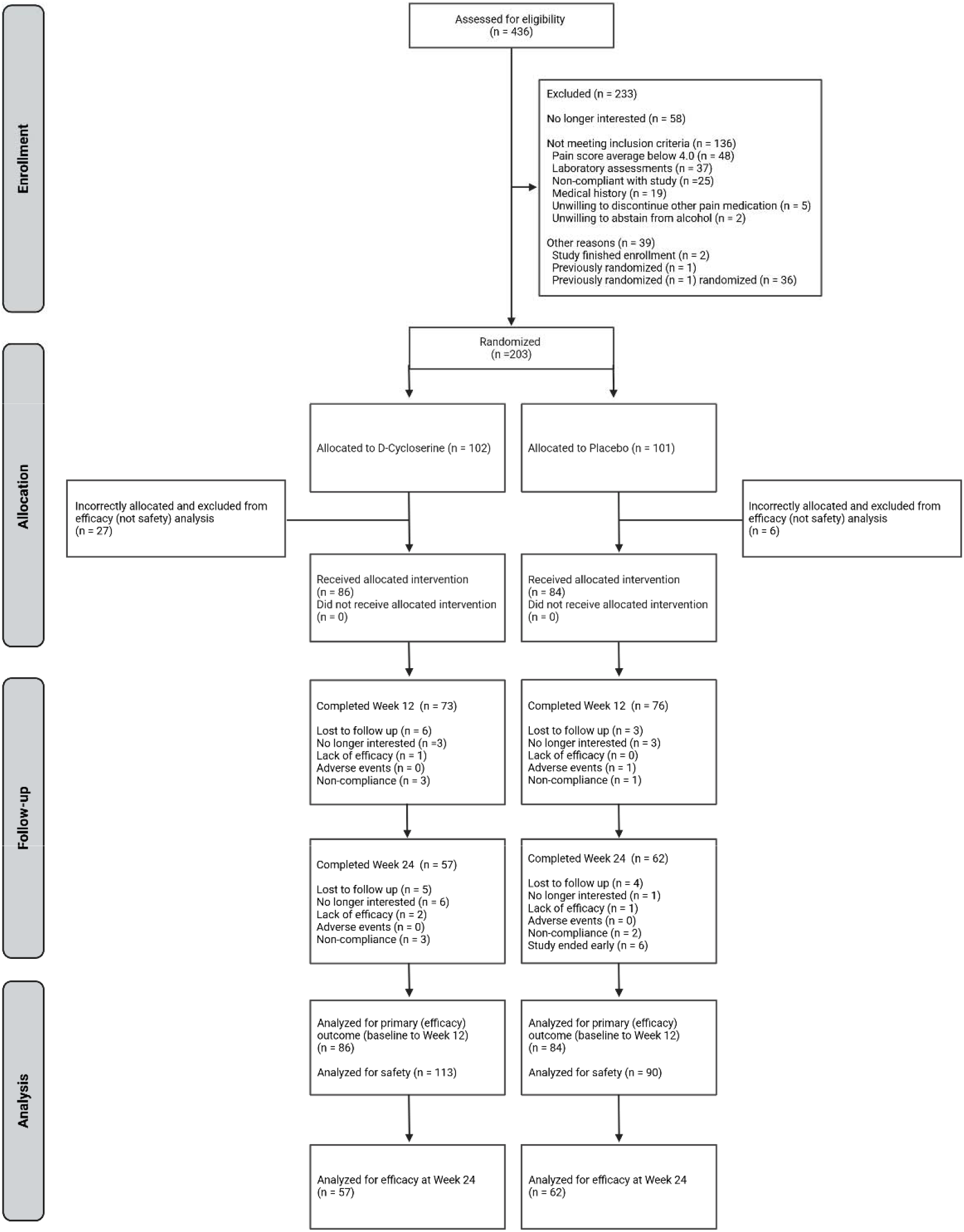
CONSORT flow diagram of participant enrollment, allocation, follow-up, and analysis. A total of 436 participants were screened for eligibility, of which 233 were excluded, primarily for not meeting inclusion criteria (n=136), followed by pain scores/demographics (n=48), laboratory assessments (n=37), non-compliance with study (n=25), medical history (n=19), unwillingness to discontinue other pain medication (n=5), and unwillingness to abstain from alcohol (n=2). A total of 203 participants were randomized, with 102 allocated to the D-cycloserine (DCS) group and 101 to the placebo group. Among all randomized, 86 participants in the DCS group and 84 in the placebo group received at least one dose of the assigned intervention and were included in the efficacy population (primary sample). All 203 randomized participants were included in the safety population. 73 participants in the DCS group and 76 in the placebo group completed the primary endpoint assessment at Week 12. By Week 24, completion rates decreased to 57 participants in the DCS group and 62 in the placebo group. Discontinuation reasons included loss to follow-up, loss of interest in participation, noncompliance, adverse events, and lack of efficacy. The primary efficacy analysis at Week 12 included 86 participants in the DCS group and 84 in the placebo group, while efficacy analyses at Week 24 included 57 participants in the DCS group and 62 in the placebo group. The final analysis included 86 DCS and 84 placebo participants for primary efficacy analysis from baseline to Week 12. The safety analysis encompassed 113 DCS and 90 placebo participants. Week 24 efficacy analysis included 57 DCS and 62 placebo participants. Notably, 27 participants in the DCS group and 6 in the placebo group were incorrectly allocated (protocol deviation identified during quality control procedures) and excluded from efficacy analysis while remaining in the safety analysis in order to maintain the integrity of the study results.

Measured baseline demographics and clinical characteristics were similar between the DCS (n=86) and placebo (n=84) groups (**Table 1**).

**Table 1.** Demographic characteristics of study participants by treatment group. Values are presented as mean ± SD for numerical variables and number (percentage) for categorical variables. BDI-II = Beck Depression Inventory – Second Version; MPQ = McGill Pain Questionnaire; ODI = Oswestry Disability Index; PANAS = Positive and Negative Affect Schedule; PCS = Pain Catastrophizing Scale; PGA = Patient Global Assessment; PGIC = Patient Global Impression of Change; SD = Standard deviation; SF-12 = 12-Item Short Form Health Survey; painDETECT = Neuropathic pain screening questionnaire.

| **Demographic and baseline patient reported measures** | **DCS (n=86)** |  | **Placebo (n=84)** |  |
| --- | --- | --- | --- | --- |
| Age (Years), Mean, SD | 56 | ±14 | 56 | ±15 |
| Gender, n (%) |  |  |  |  |
| Male | 40 | (47.6) | 45 | (53.6) |
| Female | 46 | (52.4) | 39 | (46.4) |
| Race, n (%) |  |  |  |  |
| White | 43 | (50.0) | 40 | (47.6) |
| Black or African American | 34 | (39.5) | 28 | (33.3) |
| Asian | 5 | (5.8) | 7 | (8.3) |
| Other | 4 | (4.7) | 7 | (8.3) |
| Declined to respond | 0 | (0.0) | 2 | (2.4) |
| Ethnicity, n (%) |  |  |  |  |
| Not Hispanic or Latino | 71 | (82.6) | 74 | (88.1) |
| Hispanic or Latino | 13 | (15.1) | 8 | (9.5) |
| Declined to respond | 2 | (2.3) | 2 | (2.4) |
| **Patient reported measures** |  |  |  |  |
| Baseline Pain (NRS), Mean | 6.00 | ±1.37 | 5.95 | ±1.36 |
| PANAS Positive | 34 | ±8 | 32 | ±9 |
| PANAS Negative | 17 | ±7 | 17 | ±6 |
| PDQ | 11 | ±7 | 12 | ±7 |
| BDI | 7 | ±6 | 8 | ±8 |
| ODI | 31 | ±14 | 32 | ±16 |
| SF-12 Mental Health | -0.2 | ±0.9 | -0.3 | ±1.0 |
| SF-12 Physical Health | -1.6 | ±0.8 | -1.6 | ±0.9 |
| PGA | 2.6 | ±0.6 | 2.6 | ±0.6 |
| MPQ | 14 | ±10 | 15 | ±9 |
| PCS | 13 | ±11 | 13 | ±11 |

### 3.2 Primary Outcome

The ITT analysis revealed negligible differences in pain between the DCS and placebo groups at Week 12. The adjusted treatment effect was −0.07 NRS units; the data were compatible (95% CI) with effects ranging from −0.67 to 0.53 NRS units, where negative values indicate lower pain in the DCS group. The 12-week treatment effects within each sex and the differences between the treatment effects across sexes were also highly compatible with the null hypotheses of no effect of DCS and no differences in the DCS effect between sexes (**Table 2**).

**Table 2.** Main outcome, secondary and exploratory outcomes at 12 weeks. Analyses for primary and secondary outcomes were conducted using ANCOVA adjusted for baseline pain scores. For the primary outcome (ITT pain at Week 12), blocked permutation testing was employed to maintain randomization integrity, and missing data were imputed using multivariate imputation by chained equations (MICE). Exploratory continuous outcomes (e.g., MPQ, PANAS, PDQ) were analyzed using ANCOVA, with HC3 sandwich estimators applied where appropriate to address heteroskedasticity and permutations applied when substantial departures from residual normality were present. PGA and PGIC were assessed using ordinal regression. BDI-II = Beck Depression Inventory – Second Version; MPQ = McGill Pain Questionnaire; ODI = Oswestry Disability Index; PANAS = Positive and Negative Affect Schedule; PCS = Pain Catastrophizing Scale; PGA = Patient Global Assessment; PGIC = Patient Global Impression of Change; SD = Standard deviation; SF-12 = 12-Item Short Form Health Survey; painDETECT = Neuropathic pain screening questionnaire.

| Week 12 |  |  |  |  |
| --- | --- | --- | --- | --- |
| Outcome | Estimate ± SE | 95% CI | Statistic | p-value |
| **Primary outcome** |  |  |  |  |
| NRS (ITT) | -0.07 ± 0.30 | (-0.67, 0.53) | -0.23 | 0.79 |
| Sex moderation (ITT) | -0.43 ± 0.59 | (-1.60, 0.74) | -0.73 | 0.47 |
| Effect in females (ITT) | 0.18 ± 0.42 | (-0.66, 1.01) | 0.42 | 0.67 |
| Effect in males (ITT) | -0.25 ± 0.42 | (-1.09, 0.58) | -0.60 | 0.55 |
| **Exploratory outcomes** |  |  |  |  |
| NRS (as treated) | -0.1 ± 0.3 | (-0.7, 0.4) | -0.51 | 0.61 |
| MPQ | 1.7 ± 1.2 | (-0.7, 4.1) | 1.42 | 0.16 |
| PANAS positive | -0.3 ± 1.2 | (-2.7, 2.0) | -0.26 | 0.79 |
| PANAS negative | 0.6 ± 1.0 | (-1.3, 2.5) | 0.64 | 0.52 |
| PDQ | 0.1 ± 0.8 | (-1.4, 1.6) | 0.10 | 0.92 |
| BDI | 1.8 ± 1.0 | (-0.2, 3.8) | 1.81 | 0.07 |
| ODI | 0.2 ± 2.1 | (-3.9, 4.2) | 0.08 | 0.94 |
| SF12-mental | -0.2 ± 0.1 | (-0.5, 0.1) | -1.30 | 0.20 |
| SF12-physical | 0.1 ± 0.1 | (-0.1, 0.3) | 1.19 | 0.24 |
| PGA (log OR) | -0.2 ± 0.7 | (-1.5, 1.1) | -0.31 | 0.76 |
| PGIC (log OR) | -1.3 ± 0.6 | (-2.6, -0.1) | -2.05 | 0.04 |
| PCS | 0.86 ± 1.34 | (-1.8, 3.5) | 0.64 | 0.52 |

### 3.3 Secondary outcomes

#### 3.3.1 24-week analysis

The ITT analysis revealed no significant differences in pain between the DCS and placebo groups at Week 24. The adjusted treatment effect was −0.22 NRS units; the data were compatible with effects ranging from −0.85 to 0.40 NRS units. The 24-week treatment effects within each sex and the differences between the treatment effects across sexes were also highly compatible with the null hypotheses of no effect of DCS and no differences in the DCS effect between sexes (**Table 3**). These results align with the primary Week 12 findings, indicating that the effect of daily 400 mg of DCS relative to placebo on pain ratings at 24 weeks is precisely estimated around 0.

**Table 3.** Secondary and exploratory outcomes at Week 24. Analyses for primary and secondary outcomes were conducted using ANCOVA adjusted for baseline pain scores. For the primary outcome (ITT pain at Week 24), blocked permutation testing was employed to maintain randomization integrity, and missing data were imputed using multivariate imputation by chained equations (MICE). Exploratory continuous outcomes (e.g., MPQ, PANAS, PDQ) were analyzed using ANCOVA, with HC3 sandwich estimators applied where appropriate to address heteroskedasticity and permutations applied when substantial departures from residual normality were present. PGA and PGIC were assessed using ordinal regression. BDI-II = Beck Depression Inventory – Second Version; MPQ = McGill Pain Questionnaire; ODI = Oswestry Disability Index; PANAS = Positive and Negative Affect Schedule; PCS = Pain Catastrophizing Scale; PGA = Patient Global Assessment; PGIC = Patient Global Impression of Change; SD = Standard deviation; SF-12 = 12-Item Short Form Health Survey; painDETECT = Neuropathic pain screening questionnaire.

| Week 24 |  |  |  |  |
| --- | --- | --- | --- | --- |
| Outcome | Estimate ± SE | 95% CI | Statistic | p-value |
| **Secondary outcome** |  |  |  |  |
| NRS (ITT) | -0.22 ± 0.32 | (-0.85, 0.40) | -0.70 | 0.39 |
| Sex moderation (ITT) | -0.14 ± 0.64 | (-1.41, 1.13) | -0.22 | 0.83 |
| Effect in females (ITT) | -0.12 ± 0.45 | (-1.02, 0.77) | -0.27 | 0.78 |
| Effect in males (ITT) | -0.26 ± 0.45 | (-1.15, 0.63) | -0.58 | 0.56 |
| **Exploratory outcomes** |  |  |  |  |
| NRS (as treated) | -0.4 ± 0.3 | (-1.0, 0.2) | -1.22 | 0.23 |
| MPQ | -2.2 ± 1.3 | (-4.9, 0.4) | -1.67 | 0.10 |
| PANAS positive | 0 ± 1 | (-2, 2) | 0.04 | 0.97 |
| PANAS negative | -2 ± 1 | (-4, 0) | -1.75 | 0.08 |
| PDQ | -1.1 ± 0.8 | (-2.7, 0.4) | -1.44 | 0.15 |
| BDI | -1 ± 1 | (-3, 1) | -1.18 | 0.24 |
| ODI | -1 ± 2 | (-5, 3) | -0.69 | 0.49 |
| SF12-mental | 0.1 ± 0.1 | (-0.2, 0.3) | 0.42 | 0.67 |
| SF12-physical | 0 ± 0.1 | (-0.2, 0.3) | 0.38 | 0.70 |
| PGA (log OR) | -1.1 ± 0.7 | (-2.6, 0.3) | -1.46 | 0.13 |
| PGIC (log OR) | -1.0 ± 0.6 | (-2.3, 0.2) | -1.62 | 0.10 |
| PCS | -3 ± 1 | (-5, 0) | -1.96 | 0.05 |

#### 3.3.2 Safety analysis

A total of 193 adverse events (AEs) were reported among 98 unique participants during the study, with 111 events occurring in the placebo group and 82 in the DCS group. Most events were mild in severity (n = 115, 60.2%), followed by moderate (n = 73, 37.8%), and severe (n = 5, 2.6%). Only four events (2.1%) were considered serious. Most adverse events (n = 149) were judged unrelated to the investigational product, with smaller numbers considered unlikely related (n = 23) or possibly related (n = 21). All serious adverse events were deemed unrelated to the study drug. A detailed breakdown of AEs by organ system and severity is provided in Supplementary Table 1. No unexpected safety signals or concerning patterns were identified. Overall, the treatment was well tolerated.

### 3.4 Exploratory outcomes

The as-treated analysis with imputation at Week 12 showed a nonsignificant treatment effect for DCS, with an estimated adjusted pain reduction of −0.1; the data were compatible with effects ranging from −0.7 to 0.4, excluding clinically meaningful effects in the broader (misallocated) sample. By Week 24, the adjusted effect was still modest (**Table 2**). These exploratory findings align with the primary intention-to-treat (ITT) results, reinforcing the conclusion that DCS did not demonstrate, nor were they compatible with, clinically significant analgesic efficacy compared to placebo at either timepoint.

At both Weeks 12 and 24, participants receiving active treatment reported worse on average perceived improvement compared to placebo (OR = 0.3, **Table 2**; OR = 0.4, **Table 3**); however, these estimates were imprecise. Similarly, at Week 24, PGA was lower on average in the DCS group compared to the placebo group (OR = 0.4, **Table 3**), but this difference was not precisely estimated. All other outcomes tended to have negligible effects, estimated to be close to zero.

## Discussion

In this randomized, placebo-controlled trial, we evaluated the analgesic efficacy and safety of 400 mg daily DCS for the treatment of chronic low back pain over a 24-week period. At the provided dose and duration, DCS did not demonstrate a clinically significant reduction in pain intensity compared to placebo at either the Week 12 primary endpoint or the Week 24 follow-up. These findings were consistent across both ITT and as-treated analyses. Moreover, exploratory analyses were consistent with these results. Importantly, DCS was well tolerated, with a comparable adverse event profile to that of the placebo and no unexpected safety concerns. Taken together, our results do not support the efficacy of DCS as an analgesic intervention in this population.

D-cycloserine (DCS) exhibits a biphasic, dose-dependent pharmacological profile shaped by its partial agonism at the glycine co-agonist site of NMDA receptors and regional subunit composition. At lower doses (<100 mg/day), DCS enhances NMDA receptor function and facilitates synaptic plasticity. In contrast, higher doses (>500 mg/day) saturate the receptor, competing with endogenous glycine and D-serine, resulting in functional antagonism, particularly at NR2A/NR2B subunits in cortical regions rich in glycine (e.g., the prefrontal cortex)^20, 21^. At 400 mg/day, DCS likely remains within the partial agonist range but approaches the threshold (∼500 mg) at which antagonistic effects may occur, particularly in brain regions with high endogenous glycine levels (e.g., prefrontal cortex)^20^.

This nonlinear dose–response is observed across neuropsychiatric conditions. In schizophrenia, low-dose DCS (50 mg/day) has been shown to reduce negative symptoms^22, 23^, while higher doses (100–250 mg/day) have been reported to exacerbate psychotic symptoms^24, 25^. For anxiety disorder, administering 50 mg before exposure therapy significantly enhanced fear extinction, leading to improved outcomes in both social anxiety^26^ and acrophobia^27^. Similarly, in major depressive disorder, adjunctive DCS at 100 mg/day doubled remission rates when combined with transcranial magnetic stimulation (TMS)^28^. In contrast, treatment-resistant depression (TRD) may require high-dose DCS (≥1000 mg/day) to achieve efficacy, potentially through mechanisms resembling NMDA antagonism (e.g., glutamate surge, AMPA activation, BDNF upregulation), akin to ketamine’s action^29, 30^. Thus, future studies may need to explore both higher and lower doses of DCS to identify a range where DCS may better control chronic pain.

Another important consideration regarding DCS dosing in pain treatment is the region-specific dysregulation of NMDA receptors observed in chronic pain conditions. Studies have shown that chronic pain can lead to upregulation of NR2B subunits in the spinal cord and downregulation in cortical areas^31, 32^. As a partial agonist at the glycine site of the NMDA receptor, DCS may enhance adaptive plasticity in hypoactive corticolimbic circuits but also poses a risk of overactivating hyperfunctional spinal NMDARs, potentially exacerbating nociception^33^. This dual action also underscores the critical need for precision dosing to harness DCS’s therapeutic potential while mitigating unintended amplification of maladaptive signaling.

The clinical condition under investigation, cLBP, is a complex and heterogeneous disorder in which both nociceptive and neuropathic mechanisms may contribute to symptom manifestation ^34, 35^. While our preclinical studies demonstrated the efficacy of DCS in rodent models of neuropathic pain, translating these findings to cLBP entails addressing a broader and more variable pathophysiological context. Animal models of neuropathic pain often focus on evoked hypersensitivity under well-controlled conditions, whereas cLBP in humans encompasses a multifactorial etiology involving structural degeneration, inflammation, peripheral and central sensitization, and psychological comorbidities such as depression and anxiety^34^. Despite these differences, studying DCS in cLBP offers a more ecologically valid test of its clinical potential. Importantly, the promising preclinical evidence of DCS in neuropathic pain models also underscores the need to explore its therapeutic potential in other clearly defined neuropathic pain syndromes, such as diabetic neuropathy or chemotherapy-induced peripheral neuropathy, in future clinical studies.

In summary, 400 mg/day DCS did not produce clinically significant effects on cLBP pain intensity or other secondary outcomes. The reason for DCS’s lack of efficacy in this trial remains unknown, but we surmise that it would be prudent to study DCS further in two different contexts. First, given DCS’s unique action as a partial NMDA receptor modulator with effects on synaptic plasticity, it may hold particular promise as an adjunctive therapy in contexts involving maladaptive learning, such as fear-avoidance, emotional dysregulation, or engagement in behavioral therapy. Second, DCS should be studied in other chronic pain syndromes, including well-defined neuropathic conditions such as diabetic neuropathy or chemotherapy-induced peripheral neuropathy.

## Data Availability

All data produced in the present study are available upon reasonable request to the authors

## Acknowledgememts

All authors contributed to the article and approved the submitted version. This work was funded by the ***Department of Defense Award # W81XWH-17–1-0426***. The authors report no financial or non-financial conflicts of interest in relation to the content of this manuscript.

## Notes

### Competing Interest Statement

The authors have declared no competing interest.

### Clinical Trial

NCT03535688

### Clinical Protocols

https://clinicaltrials.gov/study/NCT03535688?term=d%20cycloserine%20pain&rank=1#study-plan

### Funding Statement

This work was funded by the Department of Defense Award # W81XWH1710426

### Author Declarations

The study was approved by the Institutional Review Board of Northwestern University, which reviewed the protocol to ensure compliance with ethical standards for research involving human participants.

