## Supplemental table 1 for "D-Cycloserine for Treatment of Chronic Low Back Pain: Results from a Randomized, Double-Blind, Placebo-Controlled Clinical Trial"

|  | **Weeks 0-12** | | **Weeks 12-24** | |
| --- | --- | --- | --- | --- |
| Total AEs |  |  |  |  |
| Musculoskeletal | 23 | 19% | 17 | 29% |
| General | 39 | 32% | 8 | 14% |
| Respiratory | 10 | 8% | 5 | 9% |
| Ear | 1 | 1% | 3 | 5% |
| Immunologic | 1 | 1% | 3 | 5% |
| Nervous | 14 | 11% | 5 | 9% |
| Skin | 5 | 4% | 3 | 5% |
| Injury | 0 | 0% | 2 | 3% |
| Vascular | 4 | 3% | 2 | 3% |
| Endocrine | 0 | 0% | 2 | 3% |
| Gastrointestinal | 18 | 15% | 1 | 2% |
| Infection | 7 | 6% | 4 | 7% |
| Oral | 2 | 2% | 1 | 2% |
| Psych | 3 | 2% | 1 | 2% |
| Reproductive | 3 | 2% | 1 | 2% |
| Genitourinary | 2 | 2% | 0 | 0% |
| Eye | 3 | 2% | 0 | 0% |
| **Total AEs** | **135** |  | **58** |  |

**Supplementary Table 1.** Number and percentage of adverse events (AEs) by organ system reported during Weeks 0–12 and Weeks 12–24 of the study. Data are presented as the number of events and the percentage of total AEs in each period. Percentages are calculated based on the total number of AEs reported in the corresponding time frame (Weeks 0–12: n = 135; Weeks 12–24: n = 58).
